# White-light endoscopy versus magnifying narrow-band imaging for diagnosis of the histological subtype of gastric cancer

**DOI:** 10.1101/2020.09.21.20198846

**Authors:** Takashi Kanesaka, Noriya Uedo, Hisashi Doyama, Naohiro Yoshida, Takashi Nagahama, Kensei Otsu, Kunihisa Uchita, Koji Kojima, Tetsuya Ueo, Haruhiko Takahashi, Hiroya Ueyama, Yoichi Akazawa, Toshio Shimokawa, Kenshi Yao

**Affiliations:** Department of Gastrointestinal Oncology, Osaka International Cancer Institute, 3-1-69, Otemae, Chuo-ku, Osaka 541-8567, Japan; Department of Gastroenterology, Ishikawa Prefectural Central Hospital, 2-1, Kuratsukihigashi, Kanazawa, Ishikawa 920-8530, Japan; Department of Endoscopy, Fukuoka University Chikushi Hospital, 1-1-1, Zokumyoin, Chikushino, Fukuoka 818-8502, Japan, Japan; Department of Gastroenterology, Kochi Red Cross Hospital, 1-4-63-11, Hadaminami-machi, Kochi 780-0026, Japan; Department of Gastroenterology, Oita Red Cross Hospital, 3-2-37, Chiyo-machi, Oita 870-0033, Japan; Department of Gastroenterology, Juntendo University, School of Medicine, 2-1-1, Hongo, Bunkyo-Ku, Tokyo 113-8421, Japan; Clinical Study Support Center, Wakayama Medical University Hospital, 811-1, Kimiidera, Wakayama 641-0012, Japan

**Author notes:** **Correspondence to** Takashi Kanesaka, M.D., Department of Gastrointestinal Oncology, Osaka International Cancer Institute, 3-1-69, Otemae, Chuo-ku, Osaka 541-8567, Japan.

## Abstract

**Objective:** Distinguishing undifferentiated-type (diffuse-type) from differentiated-type (intestinal-type) cancer is crucial for determining the indication of endoscopic resection for gastric cancer. This study aimed to evaluate on-site diagnostic performance of conventional white-light endoscopy (WLE) and magnifying narrow-band imaging (M-NBI) in determining the subtype of gastric cancer.

**Design:** We conducted a multicenter prospective single-arm trial. Patients who planned to undergo treatment for histologically proven cT1 gastric cancer were recruited from six tertiary care institutions. The primary and key secondary endpoints were diagnostic accuracy and specificity, respectively. The diagnostic algorithm of WLE was based on lesion color. The M-NBI algorithm was based on the microsurface and microvascular patterns.

**Results:** A total of 208 patients were enrolled. After protocol endoscopy, 167 gastric cancers were included in the analysis. The accuracy, sensitivity, specificity, and positive likelihood ratio of WLE for undifferentiated-type cancer were 80% (95% CI 73%–86%), 69% (53%–82%), 84% (77%–90%), and 4.4 (2.8–7.0), respectively. Those of M-NBI were 82% (75%–88%), 53% (38%–68%), 93% (87%–97%), and 7.2 (3.6–14.4), respectively. There was no significant difference in accuracy between WLE and M-NBI (p=0.755), but specificity was significantly higher with M-NBI than with WLE (p=0.041). Those of M-NBI combined with WLE were 81% (74%–87%), 38% (24%–54%), 97% (92%–99%), and 11.5 (4.1–32.4), respectively.

**Conclusion:** M-NBI is more specific than WLE in distinguishing undifferentiated-type from differentiated-type gastric cancer and M-NBI combined with WLE is highly reliable (positive likelihood ratio >10).

**Trial registration number:** UMIN000032151.

**Significance of this study:** *What is already known on this subject?:*

➢ Distinguishing gastric cancer from non-cancer using endoscopy has already been validated.
➢ However, distinguishing undifferentiated-type (diffuse-type) from differentiated-type (intestinal-type) cancer is also crucial for determining the indication of endoscopic resection for gastric cancer.
➢ Several studies have proposed the characteristic findings of the subtype of gastric cancer in white-light endoscopy and magnifying narrow-band imaging.

*What are the new findings?:*

➢ Magnifying narrow-band imaging was more specific than white-light endoscopy in distinguishing undifferentiated-type gastric cancer from differentiated-type gastric cancer.
➢ The positive likelihood ratio of these combined modalities for undifferentiated-type cancer was highly reliable (>10).
➢ The present study verified the diagnostic characteristics and potential for clinical use of these two modalities.

*How might it impact on clinical practice in the foreseeable future?:*

➢ This study’s results, which included the positive likelihood ratio, suggest that optical biopsy may be introduced into the decision making of endoscopic treatment for gastric cancer.
➢ If optical biopsy based on these results is applied, the risk of surgical overtreatment is estimated to be low, leading to practical decision making.

## INTRODUCTION

The histological subtype of gastric cancer is classified into differentiated and undifferentiated types according to Nakamura’s classification [1, 2], which correspond to intestinal and diffuse types according to Lauren’s classification, respectively [3]. The indication for endoscopic resection is different between these subtypes and more restricted for the undifferentiated type compared with the differentiated type (Supplementary Figure 1) [2, 4-6]. Therefore, unlike other gastrointestinal cancers, distinguishing these subtypes is crucial for deciding the method of treatment for gastric cancer. Forceps biopsy is presently used to diagnose cancer and the subtype when a suspicious lesion is detected by gastroscopy in clinical practice.

**Figure 1.**
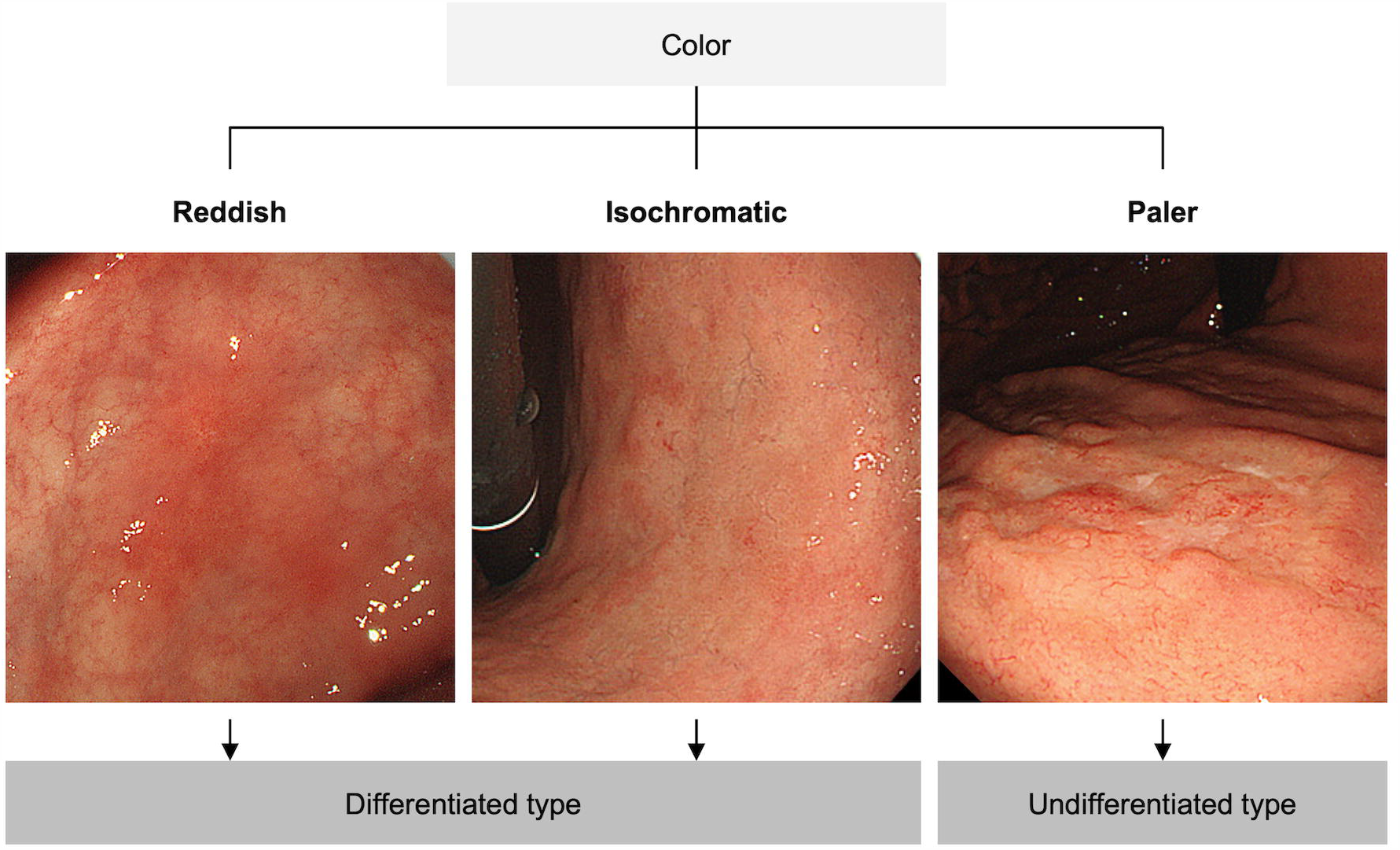
Diagnostic algorithm of white-light endoscopy for differentiating undifferentiated-type from differentiated-type gastric cancer. A paler lesion is endoscopically diagnosed as undifferentiated type, whereas a reddish or isochromatic lesion is endoscopically diagnosed as differentiated type.

Recently, magnifying narrow-band imaging (M-NBI) has been reported to be useful for the diagnosis of gastric cancer [7-10]. NBI is an image-enhanced technology, and it in combination with magnifying endoscopy allows for clear visualization of the microsurface structure and microvascular architecture of the gastric mucosa [7]. The superiority of M-NBI combined with conventional white-light endoscopy (WLE) compared with WLE alone in the diagnosis of small depressed gastric cancer has been already verified in a multicenter randomized controlled trial: accuracy increased from 64.8% to 96.6% (p < 0.001) [8]. With regard to the endoscopic diagnosis for subtypes of gastric cancer, several studies have been reported color evaluation using WLE [11, 12] and evaluation of the microsurface structure and microvascular architecture using M-NBI [13-18]. However, the diagnostic abilities of WLE and M-NBI has not been fully verified in the differentiation between differentiated-type and undifferentiated-type gastric cancer. We hypothesized that determining the subtype of gastric cancer using M-NBI could result in improved diagnostic accuracy. Therefore, this study aimed to evaluate the diagnostic performance of WLE and M-NBI in determining the subtype of gastric cancer.

## METHODS

### Study design and ethical statements

This multicenter, prospective, single-arm, phase II trial was conducted in accordance with the Declaration of Helsinki and Ethical Guidelines for Medical and Health Research Involving Human Subjects. The study protocol was approved by the Institutional Review Board of Osaka International Cancer Institute on December 22, 2017 (approval number: 1712226191) and each participating institution. This trial was registered with the University Hospital Medical Information Network Clinical Trials Registry (number: UMIN000032151) and the full study protocol can be accessed here. All authors had access to the study data and reviewed and approved the final manuscript. All participants provided written informed consent. We used the STARD checklist when writing our report [19].

### Patients

The eligibility criteria were confirmed, and informed consent was given when the patients were planned to undergo endoscopic resection or gastrectomy to treat cT1 (intramucosal or submucosal) gastric cancer in the participating institutions. If the patient agreed to participate in this trial, the preoperative endoscopic examination was undertaken according to the trial protocol.

Consecutive patients fulfilling all of the following criteria were eligible for this trial: histologically proven common-type gastric cancer (i.e., papillary adenocarcinoma, tubular adenocarcinoma, poorly differentiated adenocarcinoma [poorly differentiated tubular adenocarcinoma, solid], signet-ring cell carcinoma [poorly cohesive carcinoma], and mucinous adenocarcinoma) [20, 21]; cT1 gastric cancer; patients who planned to undergo endoscopic resection or gastrectomy in the participating institutions; and those aged ≥20 years. Patients fulfilling any of the following criteria were ineligible for this trial: risk of bleeding after biopsy, history of gastrectomy, the targeted lesion was the macroscopically elevated type, size of the targeted lesion was <5 mm, erosion or an ulcer was shown in the center of the targeted lesion, and informed consent was not obtained. Only the largest lesion was chosen in each patient for evaluation.

### Diagnostic methods

An endoscopist from the participating institutions, who was blinded to the histological subtype diagnosed in the previous examination, performed protocol endoscopy. The targeted lesion was evaluated by WLE first and then by M-NBI. Endpoints were evaluated on site according to the following WLE and M-NBI algorithms for the central part of the lesions. After completion of all diagnostic procedures, at least one biopsy specimen was obtained from the central part of the lesion.

### Evaluation with white-light endoscopy

The diagnostic algorithm of WLE used to differentiate undifferentiated-type from differentiated-type gastric cancer was based on the color of the lesions (Figure 1) [11, 12]. The location, macroscopic type, diameter, and presence of ulcer findings of the lesion were also evaluated during protocol endoscopy.

### Evaluation with magnifying narrow-band imaging

The diagnostic algorithm of M-NBI used to differentiate undifferentiated-type from differentiated-type gastric cancer was newly devised by eight expert endoscopists (K.Y., N.U., H.D., T.N., T.U., K.U., N.Y., and T.K.) on the basis of previous reports (Figure 2) [13-16].

**Figure 2.**
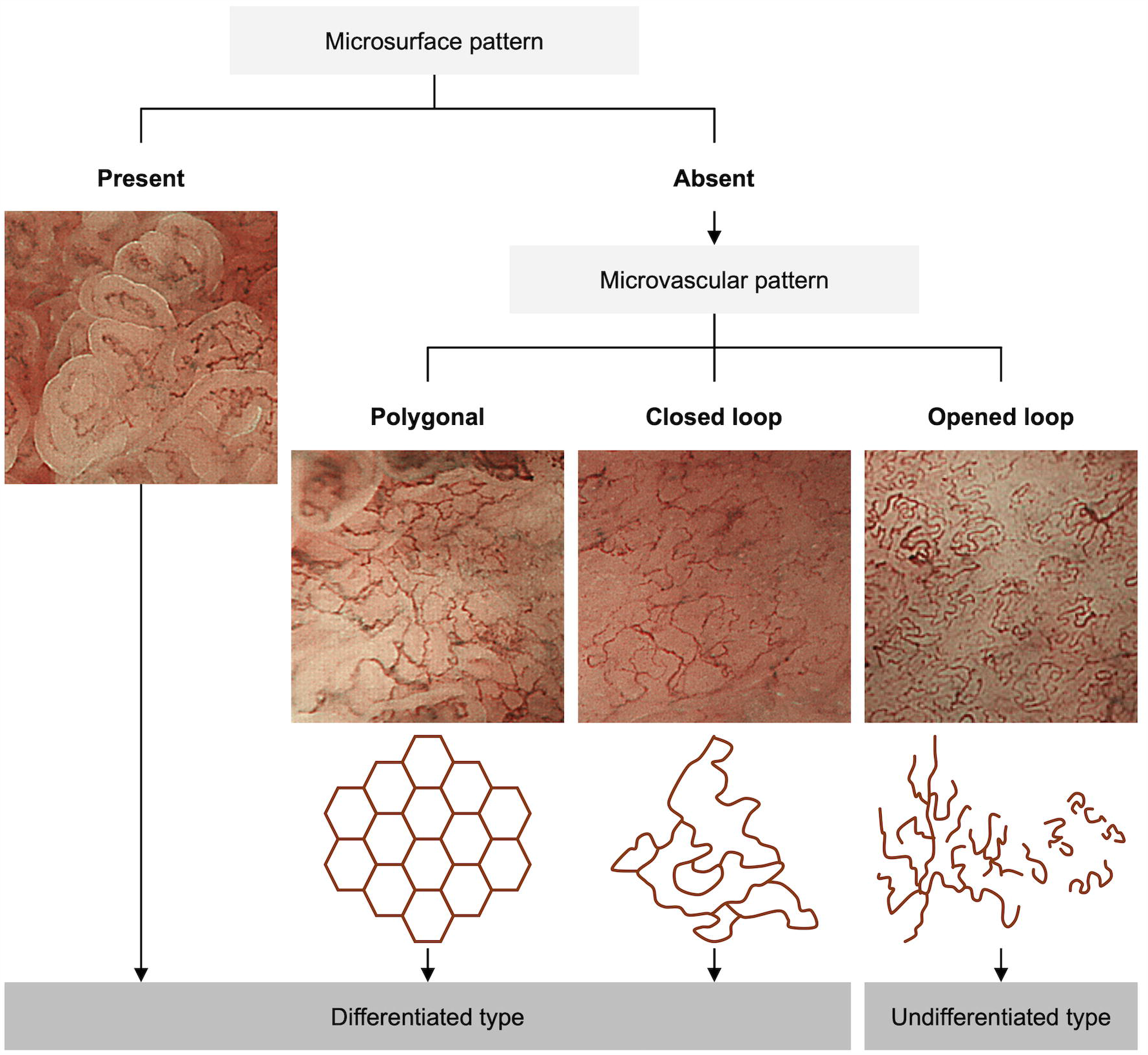
Diagnostic algorithm of magnifying narrow-band imaging for differentiating undifferentiated-type from differentiated-type gastric cancer. The lesion with a present microsurface pattern is endoscopically diagnosed as differentiated type; if the lesion shows an absent microsurface pattern, the microvascular pattern is evaluated. Polygonal or closed-loop type is endoscopically diagnosed as differentiated type, whereas opened loop type is endoscopically diagnosed as undifferentiated type.

### Histopathological diagnosis

All biopsy and resected specimens were histologically evaluated using hematoxylin and eosin staining. The pathologist was not blinded to the diagnosis based on the previous histological examination. The histological diagnosis of gastric cancer was made in accordance with the revised Vienna Classification [22]. In this trial, categories 4 (noninvasive, high-grade neoplasia) and 5 (invasive neoplasia) were classified as cancer. Categories 1 (negative for neoplasia), 2 (indefinite for neoplasia), and 3 (noninvasive, low-grade neoplasia) were classified as non-cancer. The subtype of gastric cancer was diagnosed in accordance with the Japanese classification [2, 20]. Well and moderately differentiated tubular adenocarcinoma and papillary adenocarcinoma were classified as differentiated type, and poorly differentiated adenocarcinoma and signet-ring cell carcinoma were classified as undifferentiated type. Mucinous adenocarcinoma was classified as differentiated or undifferentiated type for each case based on the degree of glandular differentiation. Mixed type was calculated as undifferentiated type.

### Outcomes

The primary and key secondary endpoints were on-site diagnostic accuracy and specificity to distinguish undifferentiated-type from differentiated-type cancer, respectively. Sensitivity, the positive likelihood ratio, and negative likelihood ratio for distinguishing undifferentiated-type from differentiated-type cancer were secondary endpoints. Adverse reactions were also investigated to evaluate the safety of M-NBI in accordance with the Common Toxicity Criteria for Adverse Events 4.03. In this trial, the central part of the lesion was endoscopically evaluated and the biopsy was taken from the same point (main analysis). In addition, diagnostic performance based on the dominant subtypes of resected specimens was calculated as a reference.

### Statistical analysis

Sample sizes were calculated to compare primary and key secondary endpoints between WLE and M-NBI. In the pilot study, 7.1% (4/56) of cT1 gastric cancers were misdiagnosed by M-NBI despite being correctly diagnosed by WLE, and 21.4% (12/56) were misdiagnosed by WLE despite being correctly diagnosed by M-NBI [23]. Using the McNemar test with a two-sided α of 0.05 and power of 0.8, 117 gastric cancers were required to compare accuracy (for the primary endpoint). Whereas, 9.8% (4/41) of differentiated-type cT1 gastric cancers were misdiagnosed by M-NBI despite being correctly diagnosed by WLE, and 24.4% (10/41) were misdiagnosed by WLE despite being correctly diagnosed by M-NBI [23]. Using the McNemar test with a two-sided α of 0.05 and power of 0.8, 132 differentiated-type cancers were required to compare accuracy between the differentiated-type cancers. Assuming that the proportion of differentiated-type cancers among cT1 gastric cancers was similar to a recent multicenter clinical trial (80.8%, 277/343) [24], 163 gastric cancers were required to compare specificity for the undifferentiated type (for the key secondary endpoint). To assess not only the primary endpoint but also the key secondary endpoint, 163 gastric cancers were required. Finally, the total sample size was set to 207 cases, considering that approximately 10% would be excluded and the false positive rate of biopsy tissue for the diagnosis of cancer would be 16.2% [25].

The baseline characteristics are summarized as median and range for continuous variables and as proportion for categorical variables. The diagnostic performance of WLE and M-NBI was assessed by accuracy, sensitivity, specificity, and the likelihood ratio. The odds ratios of matched pair data were used to summarize the differences in diagnostic performance between M-NBI and WLE. The McNemar test was used to compare the diagnostic performance between WLE and M-NBI. A p value <0.05 was considered to indicate statistical significance. All statistical analyses were conducted using R software, version 3.6.3 (R Foundation for Statistical Computing, Vienna, Austria; http://cran.r-project.org/).

### Role of the funding source

The funder had no role in this study’s design, data collection, data analysis, data interpretation, or writing of the report. The corresponding author had full access to all the data in this study and had final responsibility over the decision to submit it for publication.

## RESULTS

### Patient enrollment and background

Between September 2018 and September 2019, 208 patients were enrolled from six tertiary care institutions in Japan. Informed consent of four patients had not been stored, and one patient withdrew participation in this trial after enrollment. Among 203 patients who underwent protocol endoscopy, the procedure was completed in 192 patients by 41 endoscopists. The median number of obtained biopsy specimens was one (range, 1–2 specimens). Histological examinations of the biopsy specimen revealed that 25 lesions were not cancer. Finally, 167 cancerous lesions were included in the main analysis (Figure 3). Patient characteristics are shown in Table 1. Patients with undifferentiated-type cancer were younger than those with differentiated-type cancer (p < 0.001). Sex was similar in patients with undifferentiated-type cancer, but more men than women had differentiated-type cancer (p=0.003).

**Table 1.**
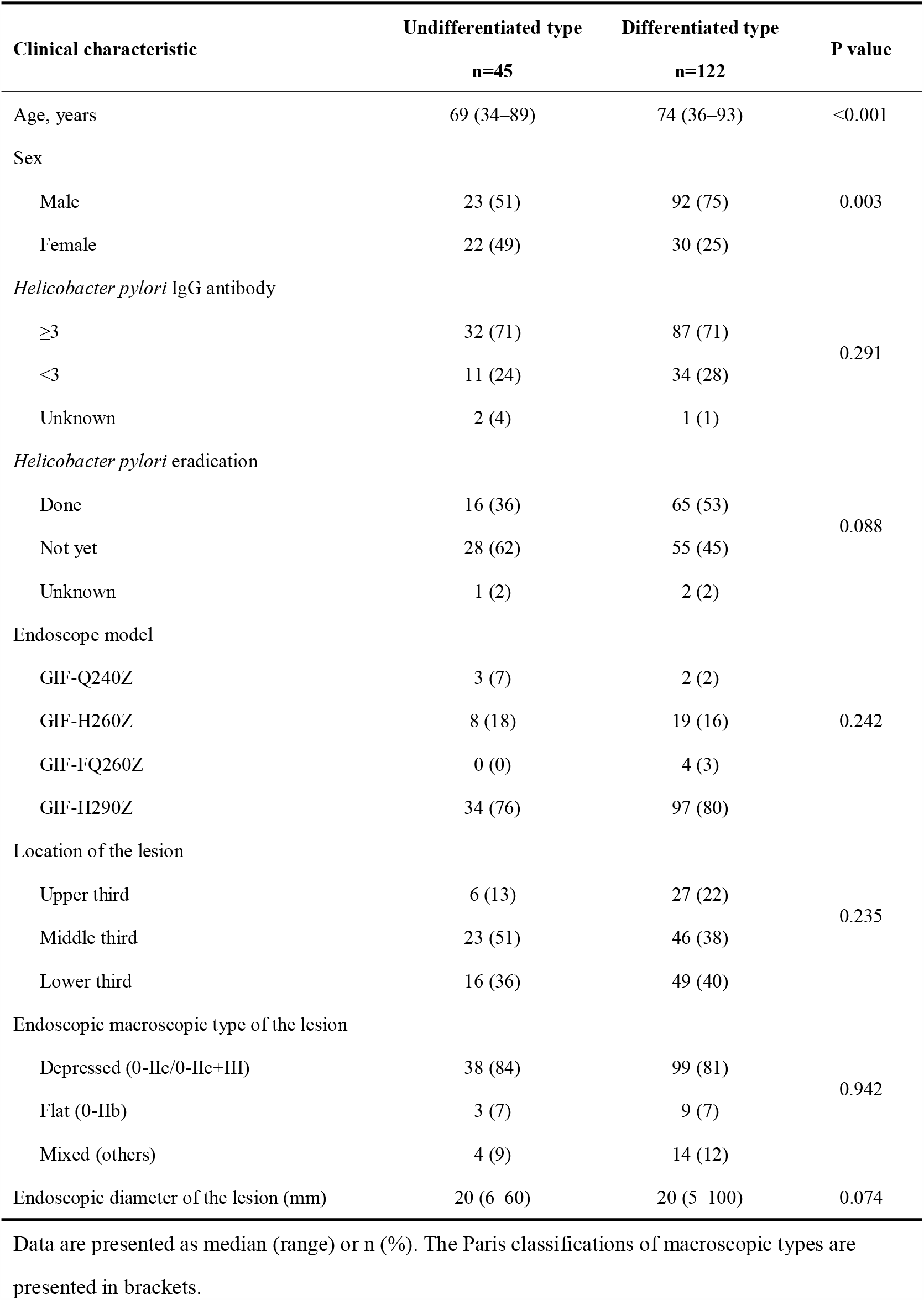
Baseline characteristics.

| Clinical characteristic | Undifferentiated type<br>n=45 | Differentiated type<br>n=122 | P value |
| --- | --- | --- | --- |
| Age, years | 69 (34–89) | 74 (36–93) | <0.001 |
| Sex |  |  |  |
| Male | 23 (51) | 92 (75) | 0.003 |
| Female | 22 (49) | 30 (25) |  |
| *Helicobacter pylori* IgG antibody |  |  |  |
| ≥3 | 32 (71) | 87 (71) | 0.291 |
| <3 | 11 (24) | 34 (28) |  |
| Unknown | 2 (4) | 1 (1) |  |
| *Helicobacter pylori* eradication |  |  |  |
| Done | 16 (36) | 65 (53) | 0.088 |
| Not yet | 28 (62) | 55 (45) |  |
| Unknown | 1 (2) | 2 (2) |  |
| Endoscope model |  |  |  |
| GIF-Q240Z | 3 (7) | 2 (2) | 0.242 |
| GIF-H260Z | 8 (18) | 19 (16) |  |
| GIF-FQ260Z | 0 (0) | 4 (3) |  |
| GIF-H290Z | 34 (76) | 97 (80) |  |
| Location of the lesion |  |  |  |
| Upper third | 6 (13) | 27 (22) | 0.235 |
| Middle third | 23 (51) | 46 (38) |  |
| Lower third | 16 (36) | 49 (40) |  |
| Endoscopic macroscopic type of the lesion |  |  |  |
| Depressed (0-IIc/0-IIc+III) | 38 (84) | 99 (81) | 0.942 |
| Flat (0-IIb) | 3 (7) | 9 (7) |  |
| Mixed (others) | 4 (9) | 14 (12) |  |
| Endoscopic diameter of the lesion (mm) | 20 (6–60) | 20 (5–100) | 0.074 |

Data are presented as median (range) or n (%). The Paris classifications of macroscopic types are presented in brackets.

**Figure 3.**
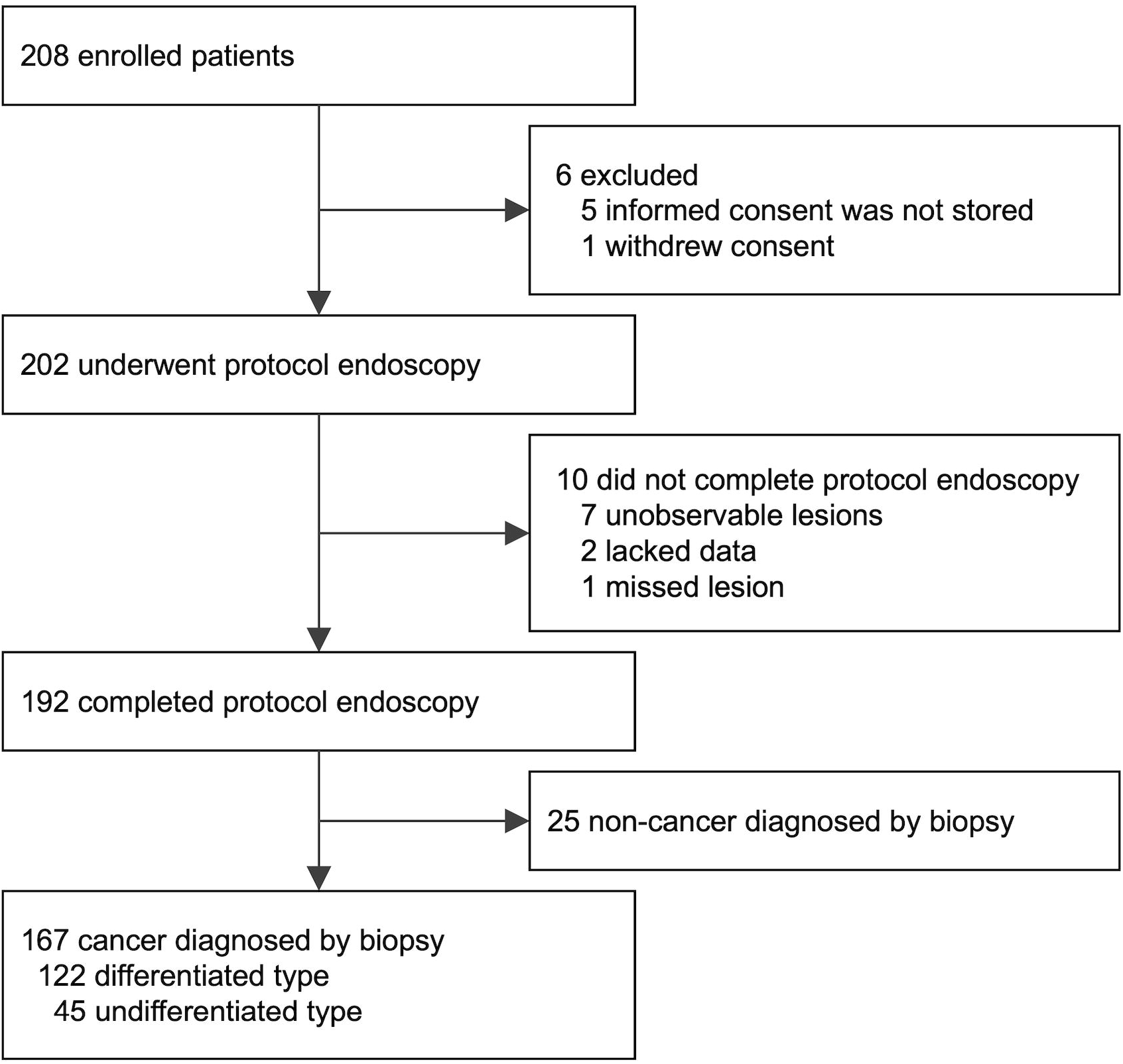
Patient flowchart.

### Diagnostic performance to distinguish subtypes of gastric cancer

In WLE, 31 undifferentiated-type cancers and 19 differentiated-type cancers were paler in color, and 14 undifferentiated-type cancers and 103 differentiated-type cancers were reddish or isochromatic in color (Table 2). The accuracy, sensitivity, and specificity for undifferentiated-type cancer were 80%, 69%, and 84%, respectively (Table 3). With M-NBI, 24 undifferentiated-type cancers and nine differentiated-type cancers showed an absent microsurface pattern and opened-loop microvessels (i.e., undifferentiated-type pattern on M-NBI), and 21 undifferentiated-type cancers and 113 differentiated-type cancers showed a present microsurface pattern and/or polygonal/closed-loop microvessels (i.e., differentiated-type pattern on M-NBI) (Table 2). The accuracy, sensitivity, and specificity for undifferentiated-type cancer were 82%, 53%, and 93%, respectively (Table 3). There was no significant difference in accuracy between WLE and M-NBI (p=0.755), but specificity was significantly higher with M-NBI than with WLE (p=0.041) (Table 4). When the lesion showed a paler color and undifferentiated-type pattern on M-NBI, the accuracy, sensitivity, specificity, and positive likelihood ratio for undifferentiated-type cancer were 81%, 38%, 97%, and 11.5, respectively (Table 3).

**Table 2.**
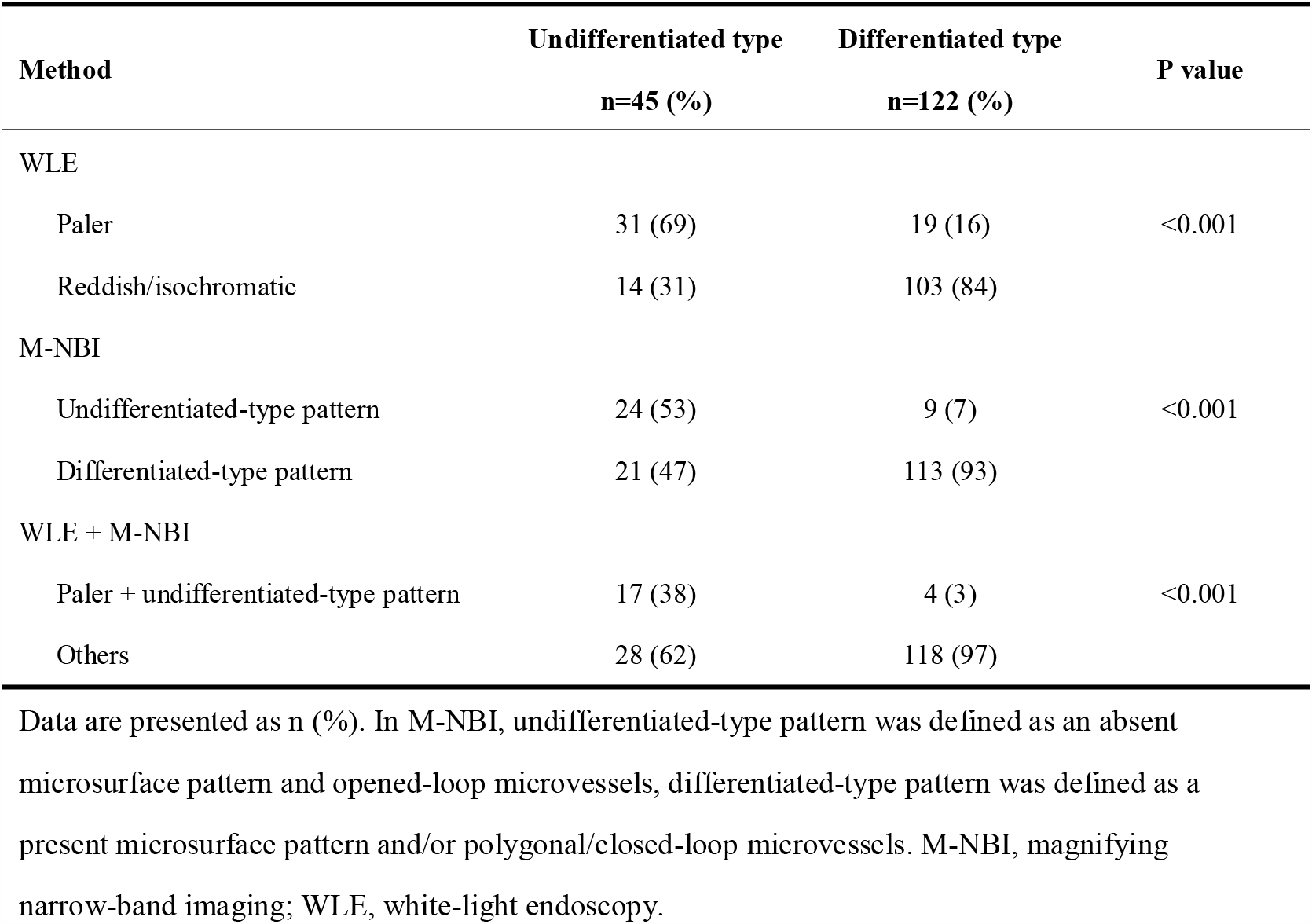
Endoscopic evaluation of the histological subtype.

| Method | Undifferentiated type | Differentiated type | P value |
| --- | --- | --- | --- |
|  | n=45 (%) | n=122 (%) |  |
| WLE |  |  |  |
| Paler | 31 (69) | 19 (16) | <0.001 |
| Reddish/isochromatic | 14 (31) | 103 (84) |  |
| M-NBI |  |  |  |
| Undifferentiated-type pattern | 24 (53) | 9 (7) | <0.001 |
| Differentiated-type pattern | 21 (47) | 113 (93) |  |
| WLE + M-NBI |  |  |  |
| Paler + undifferentiated-type pattern | 17 (38) | 4 (3) | <0.001 |
| Others | 28 (62) | 118 (97) |  |

Data are presented as n (%). In M-NBI, undifferentiated-type pattern was defined as an absent microsurface pattern and opened-loop microvessels, differentiated-type pattern was defined as a present microsurface pattern and/or polygonal/closed-loop microvessels. M-NBI, magnifying narrow-band imaging; WLE, white-light endoscopy.

**Table 3.** Diagnostic performance for undifferentiated-type gastric cancer.

| **Modality** | **Accuracy**<br>**(95% CI)** | **Sensitivity**<br>**(95% CI)** | **Specificity**<br>**(95% CI)** | **PLR**<br>**(95% CI)** | **NLR**<br>**(95% CI)** |
| --- | --- | --- | --- | --- | --- |
| WLE | 80<br>(73–86) | 69<br>(53–82) | 84<br>(77–90) | 4.4<br>(2.8–7.0) | 0.37<br>(0.24–0.57) |
| M-NBI | 82<br>(75–88) | 53<br>(38–68) | 93<br>(87–97) | 7.2<br>(3.6–14.4) | 0.50<br>(0.36–0.69) |
| WLE + M-NBI | 81<br>(74–87) | 38<br>(24–54) | 97<br>(92–99) | 11.5<br>(4.1–32.4) | 0.64<br>(0.51–0.81) |

CI, confidence interval; M-NBI, magnifying narrow-band imaging; NLR, negative likelihood ratio;
PLR, positive likelihood ratio; WLE, white-light endoscopy.

**Table 4.** Comparison of diagnostic accuracy between WLE and M-NBI for distinguishing the histological subtype of gastric cancer.

|  |  | WLE |  | Odds ratio | P value |
| --- | --- | --- | --- | --- | --- |
|  |  | Correct | Incorrect | [range] |  |
| All cT1 cancers, n=167 (%) |  |  |  |  |  |
| M-NBI | Correct | 115 | 22 | 1.2<br>[0.6–2.3] | 0.755 |
|  |  | (69) | (13) |  |  |
|  | Incorrect | 19 | 11 |  |  |
|  |  | (11) | (7) |  |  |
| Among differentiated-type cancers (i.e., specificity for the undifferentiated type), n=122 (%) |  |  |  |  |  |
| M-NBI | Correct | 98 | 15 | 3.00<br>[1.0–10.6] | 0.041 |
|  |  | (80) | (12) |  |  |
|  | Incorrect | 5 | 4 |  |  |
|  |  | (4) | (3) |  |  |

Data are presented as n (%). M-NBI, magnifying narrow-band imaging; WLE, white-light endoscopy.

Then, the diagnostic performance based on the dominant subtype of the resected specimen was also calculated. Of 192 cases who completed the protocol endoscopy, 180 lesions were diagnosed as common-type cancer (135 differentiated type and 45 undifferentiated type) based on the resected specimens (Supplementary figure 2). With WLE, 32 undifferentiated-type cancers and 21 differentiated-type cancers were paler in color, and 13 undifferentiated-type cancers and 115 differentiated-type cancers were reddish or isochromatic in color (Supplementary Table 1). The accuracy, sensitivity, and specificity of WLE for undifferentiated-type cancer were 81%, 71%, and 85%, respectively (Supplementary Table 2). With M-NBI, 25 undifferentiated-type cancers and eight differentiated-type cancers showed an undifferentiated-type pattern, and 20 undifferentiated-type cancers and 127 differentiated-type cancers showed a differentiated-type pattern (Supplementary Table 1). The accuracy, sensitivity, and specificity for undifferentiated-type cancer were 84%, 56%, and 93%, respectively (Supplementary Table 2). There was no significant difference in accuracy between WLE and M-NBI (p=0.451), but specificity was significantly higher with M-NBI than with WLE (p=0.019) (Supplementary Table 3). The accuracy, sensitivity, specificity, and positive likelihood ratio of M-NBI combined with WLE were 86%, 44%, 99%, and 60.0 (Supplementary Table 2).

### Adverse events

No adverse event occurred during and after protocol endoscopy in 203 patients.

## DISCUSSION

The purposes of forceps biopsy in the diagnosis of cT1 gastric cancer are to diagnose cancer and distinguish the histological subtype. The former using endoscopy has been already achieved [8, 9]. WLE was the worldwide standard method, and M-NBI was the more advanced imaging method for the diagnosis of gastric cancer. In this trial, the histological diagnosis corresponding to the endoscopic findings was the reference standard and was confirmed by taking biopsies from the same point. As a result, M-NBI and WLE showed high diagnostic accuracy for undifferentiated-type cancer. There was no significant difference in accuracy between these two methods. However, M-NBI showed significantly higher specificity for undifferentiated-type cancer compared with WLE. The positive likelihood ratio for undifferentiated-type cancer was sufficiently high when the lesion was diagnosed as undifferentiated type by WLE and M-NBI. Likelihood ratios above 10 and below 0.1 are considered to provide strong evidence to respectively rule in or rule out diagnoses in most circumstances [26, 27]. Therefore, M-NBI combined with WLE is a reliable diagnostic method for diagnosing the subtype of cT1 gastric cancer. This study demonstrated that the optical biopsy may be introduced into the diagnosis of gastric cancer.

As a result of the combination of WLE and M-NBI, the specificity and positive likelihood ratio for the undifferentiated type were improved but sensitivity was decreased. The combination of these methods carries a risk of resulting a false negative for the undifferentiated type because Type I error decreases and Type II error increases when the specificity increases. If the specificity is low, surgery may be indicated even for the lesion that can be cured with endoscopic submucosal dissection (ESD). Whereas, cases treated with ESD can be operated on later. According to the Japanese and Western guidelines [2, 5, 6], endoscopic resection is adapted regardless of the histological subtype if ulcerative findings were negative in T1a lesions measuring ≤2 cm. On the other hand, endoscopic resection is not adapted regardless of the histological subtype if the cancer shows massive submucosal invasion (T1b2 or deeper) or if the ulcerative findings were positive in lesions measuring >3 cm. In the present study, such 71 small lesions and 41 surgical cases were also included to reduce the selection bias. In the remaining 70 lesions, no cases are presumed to be surgically overtreated and nine (13%) cases are presumed to be required to undergo additional surgery after endoscopic resection if the endoscopic diagnosis would be applied in the decision making. Actually, two (3%) differentiated cancers were treated with surgery, and six (10%) undifferentiated-type cancers were treated with endoscopic resection. The results of this study are practical in avoiding surgical overtreatment.

Regarding the study design, we conducted a single-arm trial instead of a randomized controlled trial. WLE is generally used for the detection of suspicious gastric lesions in clinical practice [28]. M-NBI is unlikely to be used without WLE for the diagnosis of gastric cancer, because M-NBI combined with WLE was superior to M-NBI alone in distinguishing gastric cancer from non-cancer [8]. Therefore, a randomized controlled trial with a crossover of WLE and M-NBI was not suitable for the present circumstance.

Different parts of a tumor may have different histological characteristics, and it is important to define what parts of the tumor should be assessed. The main results were based on the histological examination using a biopsy specimen taken from center of the lesion to achieve the one-to-one correspondence without selection bias of the evaluation point in the lesion. However, this method might not have reflected the heterogeneity of the tumor. Moreover, all lesions that contained an undifferentiated component were considered as the undifferentiated type. Therefore, we also evaluated the diagnostic performance of the dominant subtype of resected specimens, and the results were similar.

We searched PubMed for publications on endoscopic diagnosis of histological subtype of gastric cancer published from database inception to July 31, 2020, using search terms “gastric,” “cancer,” “diagnosis,” “magnifying,” “prospective,” and (“undifferentiated” or “diffuse”), without language restrictions. Thirty-one articles were retrieved and narrowed down to three relevant prospective studies [14, 16, 18]. Two single center studies without clinical trial registries were used to evaluate the characteristic findings of the subtype of gastric cancer using M-NBI [14, 16]. In a multicenter study reported by Kishino et al [16], there was no significant difference between WLE and M-NBI with respect to the diagnostic performance for distinguishing the differentiated-type from the undifferentiated-type of gastric cancer: the accuracies of WLE and M-NBI were 96.4% (95% confidence interval [CI] 93.0%–98.4%) and 96.8% (93.6%–98.7%), respectively; whereas, sensitivities for the differentiated type, which correspond to specificity for the undifferentiated type, were 99.0% (96.5%–99.9%) and 99.5% (97.3%–100%), respectively. The proportion of the undifferentiated type among the participants of that study was very low (7% [15/221]) because only cases of endoscopic resection were included. In the guidelines, the indication for undifferentiated-type cancer is limited to cT1a (intramucosal) with a lesion size ≤2 cm, whereas the indication for differentiated-type cancer has no restriction for lesion size if it is cT1a without ulcer findings [2, 5, 6]. A strength of our study was that cases of endoscopic resection and surgical cases were included to reduce selection bias.

Why is undifferentiated-type gastric cancer paler in color and differentiated-type gastric cancer reddish or isochromatic in color? Yao et al. quantitatively measured the hemoglobin volume in gastric mucosa to answer this question [12]. Among differentiated-type gastric cancers, mucosal vascularity was higher or the same in the lesions compared with the surrounding mucosa. However, among undifferentiated-type gastric cancers, mucosal vascularity was lower in the lesions compared with the surrounding mucosa. Distinguishing the subtype of gastric cancer with M-NBI is based on the evaluation of the presence or absence of cancerous ductal formation. In M-NBI, marginal crypt epithelium of gastric mucosa is visible unless the crypt is shallow or the intervening part is short [29]. Visualization of the microsurface structure in the cancerous part means the presence of cancerous ducts [30], namely the feature of differentiated-type cancer. Thus, the presence of the cancerous duct can be speculated from the shape of the microvascular architecture. According to the evaluation of the microvascular architecture of gastric cancer using the CD31-immunostained histopathological examination, microvessels were present around the cancerous ducts in the surface mucosa of differentiated-type cancer (polygonal/closed-loop microvessels) [13]. Therefore, WLE and NBI discriminate between differentiated-type and undifferentiated-type cancers by different principles.

Our study has some limitations. First, only patients with histologically proven cT1 gastric cancer were recruited in this trial. Second, only color evaluation was included in the algorithm of WLE. Color had been demonstrated as an associated factor with the histological subtype of gastric cancer in previous retrospective studies [11, 12]. Whereas, the other factors had been already evaluated in the post hoc analysis of a multicenter prospective trial, and macroscopically elevated-type lesion was significantly associated with the differentiated-type gastric cancer with a very high positive likelihood ratio (15.7 [95% CI 2.2–110.8]) [31]. Third, as a result, macroscopically elevated-type lesions were excluded.

In conclusion, M-NBI combined with WLE is a reliable method for diagnosing the subtype of cT1 gastric cancer. Based on this evidence, the endoscopic diagnosis can replace the biopsy diagnosis in the diagnosis of cT1 gastric cancer.

## Data Availability

Data are available in the links below.

https://www.umin.ac.jp/icdr/index.html

## Acknowledgements

This work was supported by the Yasuda Medical Foundation. We would like to thank the following. Implementing medical institutions: Ryu Ishihara, Yoji Takeuchi, Koji Higashino, Noriko Matsuura (Osaka International Cancer Institute, Osaka), Ryosuke Ohta, Yasuhito Takeda, Kenichi Takemura, Kunihiro Tsuji, Shigetsugu Tsuji, Kei Tominaga, Hisashi Doyama, Hiroyoshi Nakanishi, Kazuhiro Matsunaga, Shinya Yamada, Naohiro Yoshida (Ishikawa Prefectural Central Hospital, Ishikawa), Satoshi Ishikawa, Hiroshi Ishihara, Kentaro Imamura, Kensei Ohtsu, Yoichiro Ono, Takao Kanemitsu, Kenjiro Kuhara, Toshiki Kojima, Haruhiko Takahashi, Takashi Nagahama, Shoko Fujiwara, Takahiro Beppu, Kenshi Yao (Fukuoka University Chikushi Hospital, Fukuoka), Tetsuya Ueo (Oita Red Cross Hospital, Oita), Kunihisa Uchita (Kochi Red Cross Hospital, Kochi); data center: Keiko Shindo (Osaka International Cancer Institute, Osaka); Effectiveness and Safety Evaluation Committee: Hideki Ishikawa (Kyoto Prefectural University of Medicine, Kyoto), Manabu Muto (Kyoto University, Kyoto); monitoring: Yasushi Yamasaki (Okayama University Hospital, Okayama); and English language editing: Editage (www.editage.com).

## Contributors

TK, NU, HD, NY, TN, KU, TU, and KY contributed to the study concept and design. TK, NU, HD, NY, KO, KU, KK, TU, HT, HU, YA, and KY collected data. TK analyzed and interpreted data, wrote the manuscript, and obtained funding. TS contributed to the statistical analysis. NU and KY supervised TK. All authors approved the final version of the article.

## Funding

The Yasuda Medical Foundation.

## Disclaimer

The funding source had no role in the conduct of the study; the collection, management, analysis or interpretation of the data; or in the preparation, review or approval of the manuscript.

## Competing interests

We declare no competing interests.

## Patient and public involvement

Patients and/or the public were not involved in the design, or conduct, or reporting, or dissemination plans of this research.

## Patient consent for publication

Not required.

## Provenance and peer review

Not commissioned; externally peer reviewed.

## Data availability statement

Data are available on reasonable request. The data that support the findings of this study have been deposited in UMIN (https://www.umin.ac.jp/icdr/index.html), and the data are available from the corresponding author, TK, on reasonable request.

## Notes

### Competing Interest Statement

The authors have declared no competing interest.

### Clinical Trial

UMIN000032151

### Clinical Protocols

https://www.umin.ac.jp/icdr/index.html

### Author Declarations

The study protocol was approved by the institutional Review Board of Osaka International Cancer Institute, institutional Review Board of Ishikawa Prefectural Central Hospital, institutional Review Board of Fukuoka University Chikushi Hospital, institutional Review Board of Kochi Red Cross Hospital, institutional Review Board of Oita Red Cross Hospital, and institutional Review Board of Juntendo University.

